# Developing a Tiered Machine Learning Alert System for Real-Time Suicide Risk Detection in a Digital Mental Health Setting

**DOI:** 10.64898/2026.03.26.26349452

**Authors:** Macayla L. Donegan, Agrima Srivastava, Emily Peake, Mackenzie Swirbul, Alby Ungashe, Michael Rodio, Nir Tal, Gil Margolin, Nikole Benders-Hadi, Aarthi Padmanabhan

**Affiliations:** Talkspace, LLC P. O. Box 659 Portsmouth, NH 03802

## Abstract

The goal of this work was to leverage a large corpus of text based psychotherapy data to create novel machine learning algorithms that can identify suicide risk in asynchronous text therapy. Advances in the field of natural language processing and machine learning have allowed us to include novel data sources as well as use encoding models that can represent context.

Our models utilize advanced natural language processing techniques, including fine-tuned transformer models like RoBERTa, to classify risk. Subsequent model versions incorporated non-text data such as demographic features and census-derived social determinants of health to improve equitable and culturally responsive risk assessment, as well as multiclass models that can identify tiered levels of risk.

All new models demonstrated significant improvements over our previous model. Our final version, a multiclass model, provides a tiered system that classifies risk as “no risk,” “moderate,” or “severe” (weighted F1 of 0.85). This tiered approach enhances clinical utility by allowing providers to quickly prioritize the most urgent cases, ensuring a more accurate and timely intervention for clients in need.

**Author Summary:** Suicide is a major public health concern, and traditional methods for assessing risk in clinical settings have serious limitations, often failing to capture risk in real time. To address this, we developed a series of new machine learning models to automatically and accurately detect suicide risk from the text of therapy messages. By training these models on a large, unique dataset of de-identified clinical transcripts, we were able to move beyond simple keyword spotting to a more contextual interpretation of a client’s language. The resulting models showed vast improvements over our previous published model. This is critical both for catching as much risk as possible, and for reducing “alert fatigue” for our clinicians by reducing the number of false alarms raised by the model. Furthermore, our final model, v3.0, introduced a tiered system that classifies text as “no risk,” “moderate,” or “severe.” This allows clinicians to quickly prioritize the most urgent cases, ensuring a more accurate, equitable, and timely intervention for the clients who need it most.

---

Suicide remains a leading cause of death in the United States, representing a critical priority for national public health.^1^ In 2023 alone, 12.8 million people in the United States seriously considered suicide, 3.7 million made a plan, and 1.5 million attempted suicide.^1^ Despite the high public health burden, traditional self-report suicide assessments have clear limitations, including patient underreporting, recall bias, and delayed detection by clinicians,^2^ underscoring the need for innovative tools that can identify risk in real time.

To address these limitations, recent approaches increasingly leverage machine learning and natural language processing (NLP) to detect early signals of suicide risk from digital communication. The landscape of risk detection is evolving rapidly, with artificial intelligence (AI) systems being trained to identify suicidal and self-harm risk through linguistic patterns, behavioral markers, and other digital traces.^3^ A critical emerging direction involves direct integration of risk-detection models into clinical workflows, such as therapist dashboards and automated triage systems. These implementations allow algorithmic alerts to trigger rapid human review, facilitating timely intervention.^4^

In 2019, Talkspace, a behavioral telehealth platform, launched its original suicide risk detection model (v1.0), which analyzes asynchronous text exchanges between therapy providers and clients to detect language patterns associate with elevated suicide risk.^5^ The primary objective of the previously published (v1.0) algorithm was to narrow the temporal gap between a client’s indication of suicide ideation (SI) and a provider’s intervention. By flagging potentially risky messages for immediate clinical review, the system empowers providers to exercise clinical judgment and intervene more promptly, potentially mitigating acute crises.^4^

Since the deployment of v1.0, rapid advancements in machine learning techniques have introduced more sophisticated architectures such as transformer models for text encoding and neural network classifiers that may improve the ability to accurately predict risk. These advances have made it possible to move beyond keyword detection toward more contextual interpretations of language.^6^ Unlike earlier methods, modern transformer models can distinguish between linguistically similar but semantically distinct statements such as “I’m thinking about ending it” when it refers to non-suicidal situations like ending a relationship versus suicidal intent. With sufficient contextual detail and by capturing these nuances, risk classification can be informed by meaning and intent rather than the presence of specific high-risk terms.

In the current study, we developed and validated a series of machine learning models designed to automatically identify suicide risk levels within psychotherapy transcripts. Our first iteration of the updated SI model, v2.0, utilizes state-of-the-art machine learning techniques, to assess asynchronous text, and transcribed voice, and video messages in an updated corpus of data. This expansion of data types from the previous v1.0 ensures the model remains representative of an increasingly diverse clinical population accessing mental health care through virtual platforms. Since the deployment of v1.0, the inclusion of a larger cohort, provides the necessary volume to capture heterogeneity in linguistic nuances and inter-individual differences in communication. Therefore, the model was able to capture overall language shifts^7^ and adapt to how people access care over time.^8^

The following version (v2.1) sought to enhance predictive power by incorporating contextual information. Version 2.1 integrated demographic data and census derived social determinants of health (SDOH), as factors such as age, gender, race/ethnicity, socioeconomic status, and neighborhood context are known to influence both baseline risk and linguistic expression of distress.^9,10^

Finally, version 3.0 introduced a tiered detection system to distinguish between different levels of risk using message data only. This stratification aims to enhance clinical utility by providing a prioritized triage system that alerts and identifies the risk as either no, intermediate, or high acute risk, which may decrease alert fatigue and ensure that the most acute cases receive the most rapid escalation. Here we detail the development and validation of these iterative models, evaluating their performance to provide a scalable system for real-time suicide risk detection in clinical digital settings.

## Materials and Methods

### Participants and Setting

We analyzed a corpus of 50,000 psychotherapy transcripts derived from messaging based psychotherapy on the Talkspace platform between January 1, 2020 and July 1, 2024. This corpus includes transcripts from asynchronous text, voice, and video messages. These data involve licensed US-based therapists providing care to individuals (‘clients’) via a secure digital interface on the Talkspace platform. Notably, Talkspace’s models are trained using a large dataset of de-identified clinical transcripts, which are historically difficult to access in the broader field due to ethical and privacy-related barriers. Talkspace, with its volume of clinical data, is uniquely equipped to both train NLP models and subsequently test and/or apply them. Most suicide-risk machine learning models depend on public data sources such as social media, which lack clinical context and do not reflect real therapeutic communication.^11^

The therapeutic model centered on flexible asynchronous messaging; clients could message their providers at their convenience through the platform, while therapists were instructed to maintain a consistent presence by responding at least once every 24 hours during the work week. All sessions adhered to established ethical guidelines, including the provision of referrals for individuals requiring intensive psychiatric care.

### Design and Procedures

The current study implemented a Large Language Model (LLM) based labeling system, guided by the SI coding framework established in v1.0^5^ which served as a foundation for our instruction set. Labeling was performed using the GPT4.1 model, which served as a foundation for our instruction set. The LLM-labeled test dataset was independently reviewed by multiple human annotators, with final reference labels derived using a majority vote consensus.

To further assess labeling reliability, a secondary LLM was employed as an independent evaluator (“LLM as a judge”) for all messages initially labeled as SI-3 or higher. The judge model reviewed each risky message and assessed whether the assigned label was justified by the text content or not by generating a confidence score on a 10-point scale, reflecting its certainty in the alignment between the assigned label and the message content. This secondary step was designed to act as an automated quality control mechanism for high risk cases, which are clinically important but often more semantically nuanced and susceptible to labeling noise. Only messages for which the judge model indicated high confidence (confidence >=8) in label correctness were retained for subsequent analysis (moderate risk = 3686 vignettes, severe risk = 281, no risk subsampled to 10000 vignettes for modeling).

### Model evaluation metrics

Model performance was assessed using the following metrics:

#### F1(weighted)

The F1-score is defined as the harmonic mean of recall and precision, which can be interpreted as the average of the two proportion-based metrics to give an overall measure of model accuracy. The weighted F1 score is a metric for multi-class models that calculates the F1 score for each individual class and then averages them together, giving more “weight” to classes with more examples in the dataset.

#### F2

Similar to F1, but higher weight is given to recall, which is useful in scenarios where false negatives are more costly than false positives, such as SI detection.

#### Precision

Precision is the proportion of true positives among the population of observations predicted as the positive class.

#### Recall

Recall is defined as the proportion of true positives among the observations that had a ground truth label of risk.

#### Receiver operating characteristic area under the curve (ROC AUC)

The ROC AUC is a scalar performance metric representing the probability that a classifier will rank a randomly chosen positive instance higher than a negative one, measuring the model’s ability to discriminate between classes across all possible classification thresholds.

#### Accuracy

Accuracy is the percentage of predicted labels that are correct.

### Model v2.0 Training

In the first model (v2.0), we used messages labeled by the LLM system to train classification models to determine whether text contained risk for SI. We concatenated the risk-labeled message with four prior client messages to create five-message vignettes to incorporate context. We tokenized these vignettes using the RoBERTa^12^ and ELECTRA^13^ tokenizers and extracted additional text features (Table 2) from these vignettes, such as readability and number of words related to death.

**Table 1.**
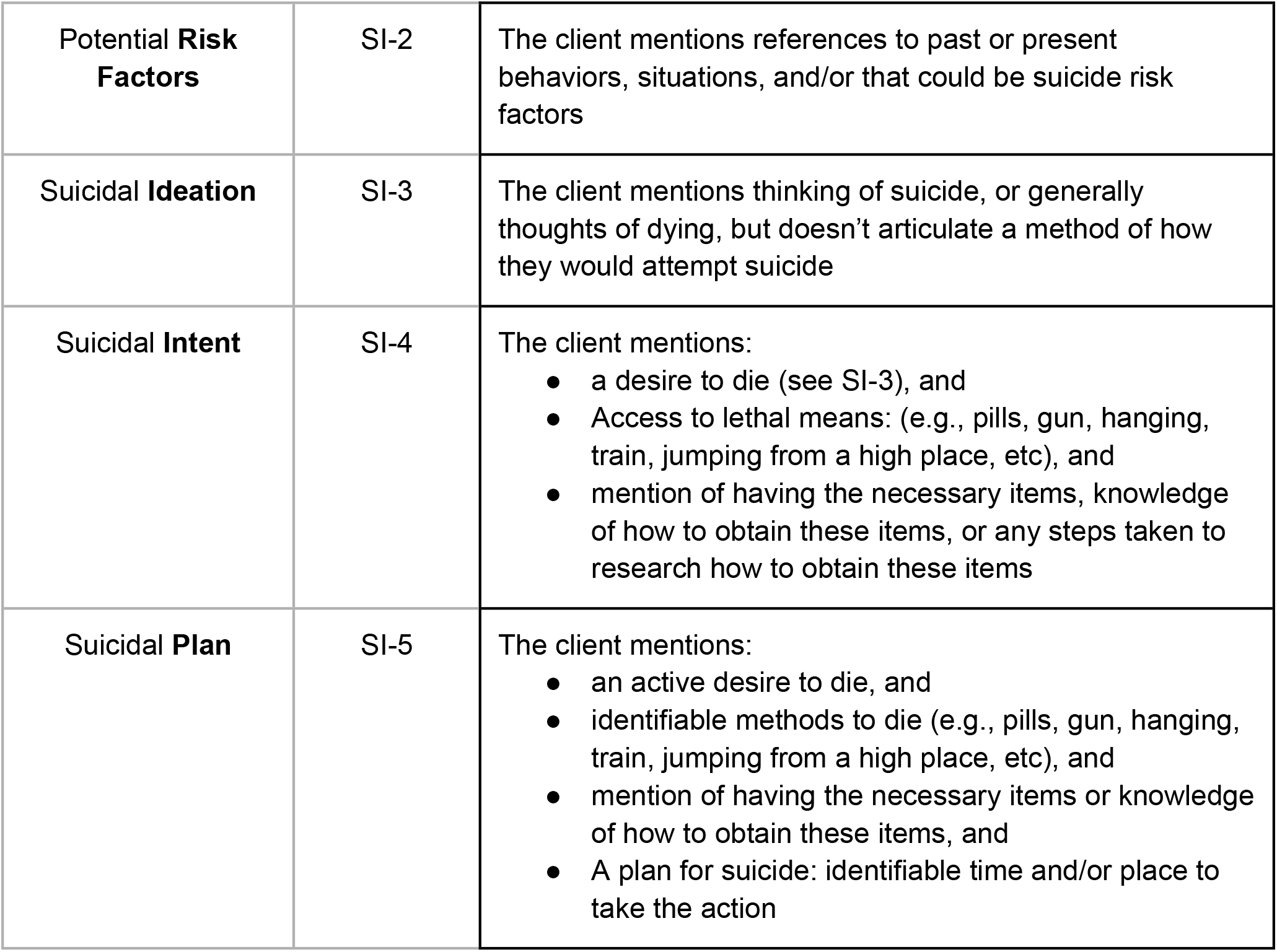
Risk level coding framework.

|  |  |  |
| --- | --- | --- |
| Potential **Risk Factors** | SI-2 | The client mentions references to past or present behaviors, situations, and/or that could be suicide risk factors |
| Suicidal **Ideation** | SI-3 | The client mentions thinking of suicide, or generally thoughts of dying, but doesn't articulate a method of how they would attempt suicide |
| Suicidal **Intent** | SI-4 | The client mentions:<br>• a desire to die (see SI-3), and<br>• Access to lethal means: (e.g., pills, gun, hanging, train, jumping from a high place, etc), and<br>• mention of having the necessary items, knowledge of how to obtain these items, or any steps taken to research how to obtain these items |
| Suicidal **Plan** | SI-5 | The client mentions:<br>• an active desire to die, and<br>• identifiable methods to die (e.g., pills, gun, hanging, train, jumping from a high place, etc), and<br>• mention of having the necessary items or knowledge of how to obtain these items, and<br>• A plan for suicide: identifiable time and/or place to take the action |

**Table 2:**
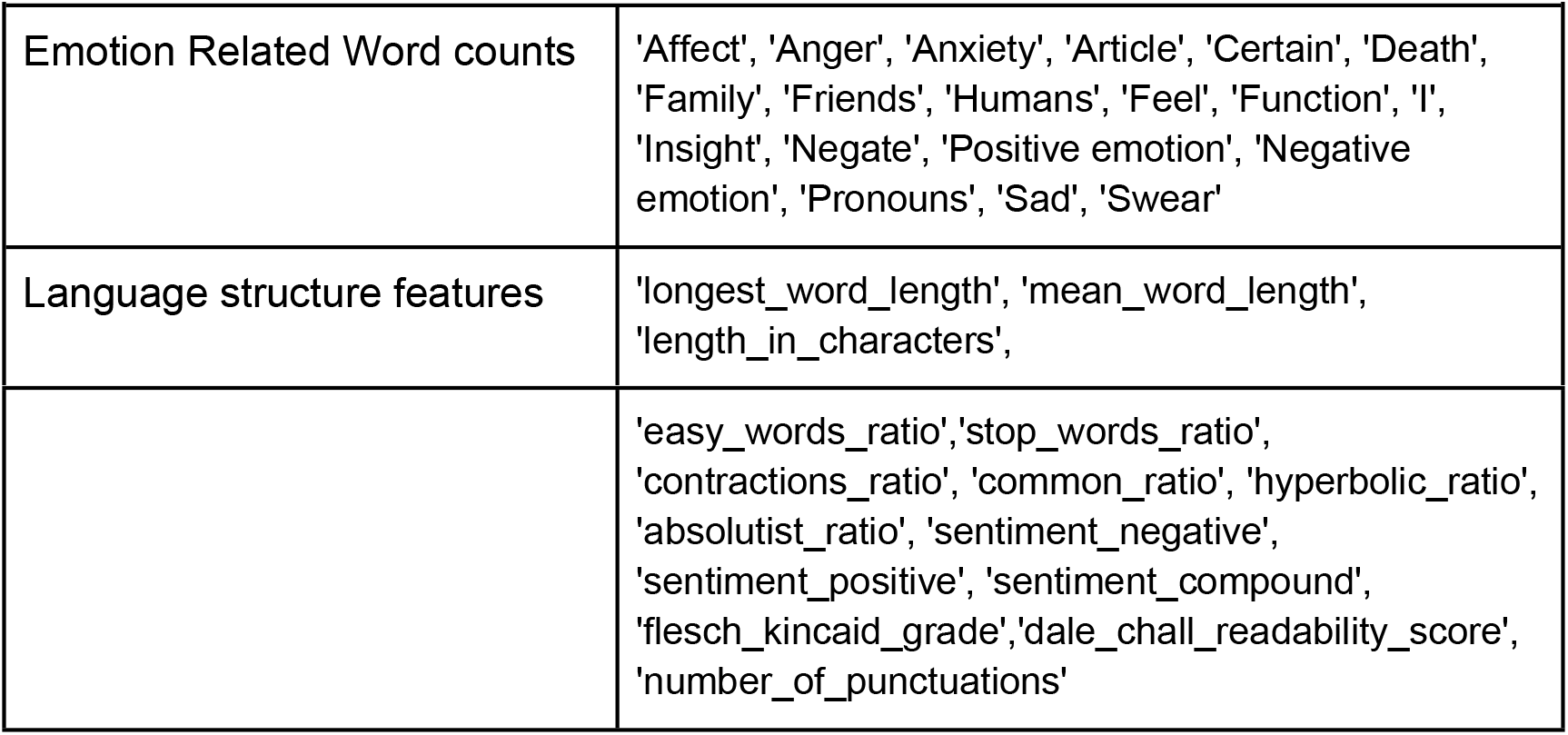
Features.

We used a 75/25 (n = 10467/3488) split for training and testing data and trained a number of models using these features. These included a logistic regression (lr), linear support vector machine (svm), naive bayes (nb), k-nearest neighbors (knn), random forest (rf), xgboost (xgb), and several neural networks using different combinations of extracted text features and text embeddings. For model v2.0 we treated this as a binary classification problem, training the models to classify “risk” (SI-3 or above) vs “no risk” (SI-2 or no risk indicated). In addition to using shallow neural networks on the above features, we also used fine-tuned foundation models for text classification, including RoBERTa^12^ and ELECTRA.^13^ We again used a 75/25 (n = 10467/3488) train/test split of the data, with the test set split into a validation dataset for tuning weights in the models. We used GridSearchCV^14^ and Optuna^15^ to determine the best hyperparameters for each model. The models were then evaluated on the whole test dataset.

### Model v2.1 Training

Previous research has demonstrated that demographics and social determinants of health may be predictive of risk for SI.^16-24^ Thus, version 2.1 incorporated demographic information, census-derived social determinants of health (SDOH) information based on ZIP code-level data, and participants’ scores on their most recent PHQ-9 and the GAD-7 collected prior to the labeled messages (Table 3). Missing data was input as -1 to the model, as the absence of survey data may itself be a predictor of risk. To combine text and tabular features, we leveraged Autogluon’s multimodal predictor.^25^ Again, we used a 75/25 (n = 10467/3488) split for training and testing data and trained a number of models using these features.

**Table 3.**
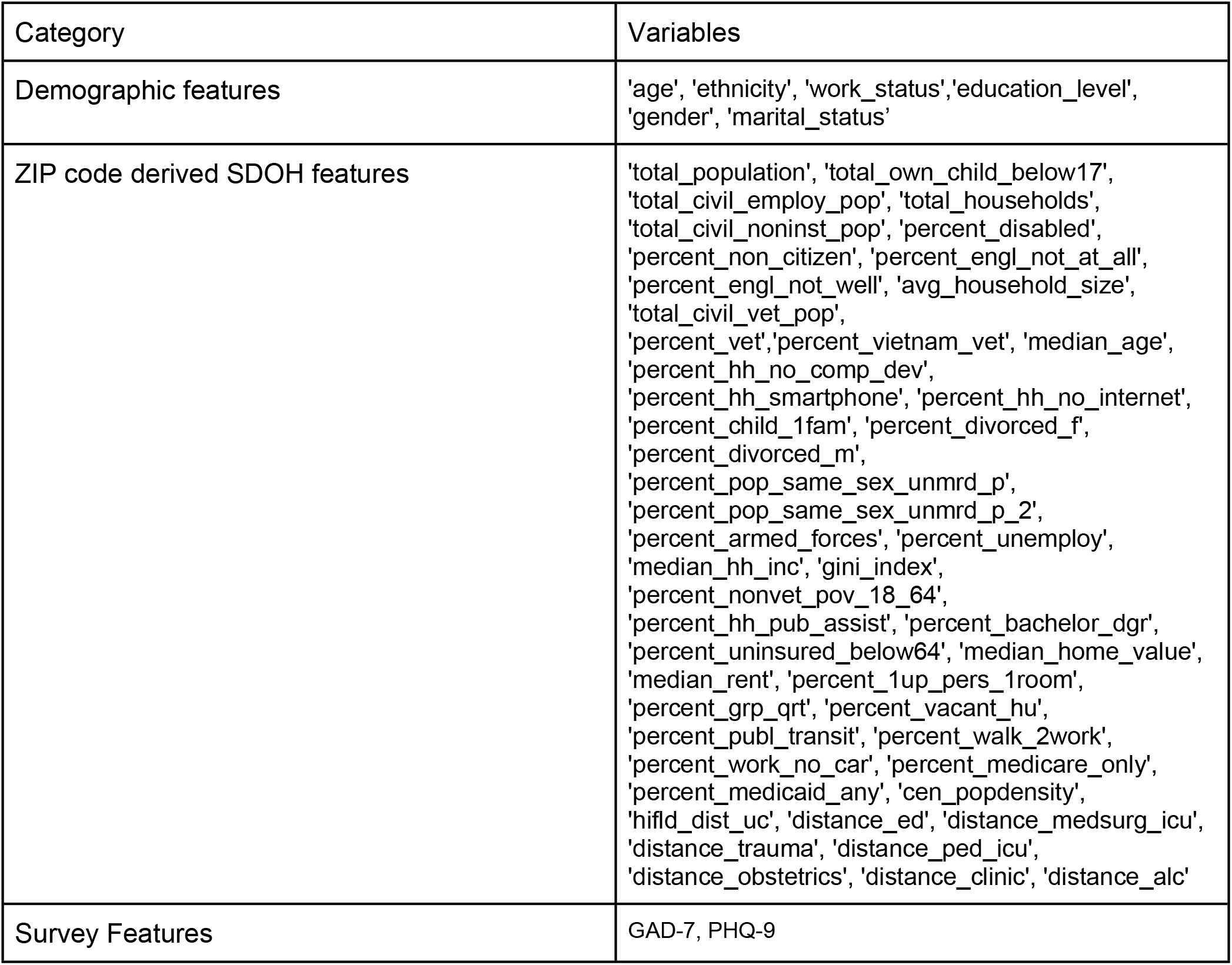
Non-text features.

| Category | Variables |
| --- | --- |
| Demographic features | 'age', 'ethnicity', 'work_status', 'education_level', 'gender', 'marital_status' |
| ZIP code derived SDOH features | 'total_population', 'total_own_child_below17', 'total_civil_employ_pop', 'total_households', 'total_civil_noninst_pop', 'percent_disabled', 'percent_non_citizen', 'percent_engl_not_at_all', 'percent_engl_not_well', 'avg_household_size', 'total_civil_vet_pop', 'percent_vet', 'percent_vietnam_vet', 'median_age', 'percent_hh_no_comp_dev', 'percent_hh_smartphone', 'percent_hh_no_internet', 'percent_child_1fam', 'percent_divorced_f', 'percent_divorced_m', 'percent_pop_same_sex_unmrd_p', 'percent_pop_same_sex_unmrd_p_2', 'percent_armed_forces', 'percent_unemploy', 'median_hh_inc', 'gini_index', 'percent_nonvet_pov_18_64', 'percent_hh_pub_assist', 'percent_bachelor_dgr', 'percent_uninsured_below64', 'median_home_value', 'median_rent', 'percent_1up_pers_1room', 'percent_grp_qrt', 'percent_vacant_hu', 'percent_publ_transit', 'percent_walk_2work', 'percent_work_no_car', 'percent_medicare_only', 'percent_medicaid_any', 'cen_popdensity', 'hifld_dist_uc', 'distance_ed', 'distance_medsurg_icu', 'distance_trauma', 'distance_ped_icu', 'distance_obstetrics', 'distance_clinic', 'distance_alc' |
| Survey Features | GAD-7, PHQ-9 |

### Model v3.0 Training

Finally, as risk severity is an important component of effective triage and crisis management, we created a multiclass model to differentiate between tiered levels of risk. We defined the highest level of risk as suicidal method or plan (SI-4, SI-5), moderate risk as suicidal ideation (SI-3), and no risk when no immediate risk indication was present (SI-1, SI-2, not labeled). Again, we used a 75/25 split (n = 10467/3488) for training and testing data and trained a number of models using these features.

### Ethical Considerations

All data were collected as part of ongoing business operations. Data were deidentified prior to analysis, and the use of patient data was reviewed and deemed exempt by an institutional review board. Clients and providers gave consent for the use of their deidentified data for research and product development purposes by accepting the platform’s Privacy Policy and Terms of Use at account creation. Model development was based on deidentified therapy transcripts which remained in an encrypted Talkspace database and were not accessed by the researchers at any time.

## Results

### Client Characteristics

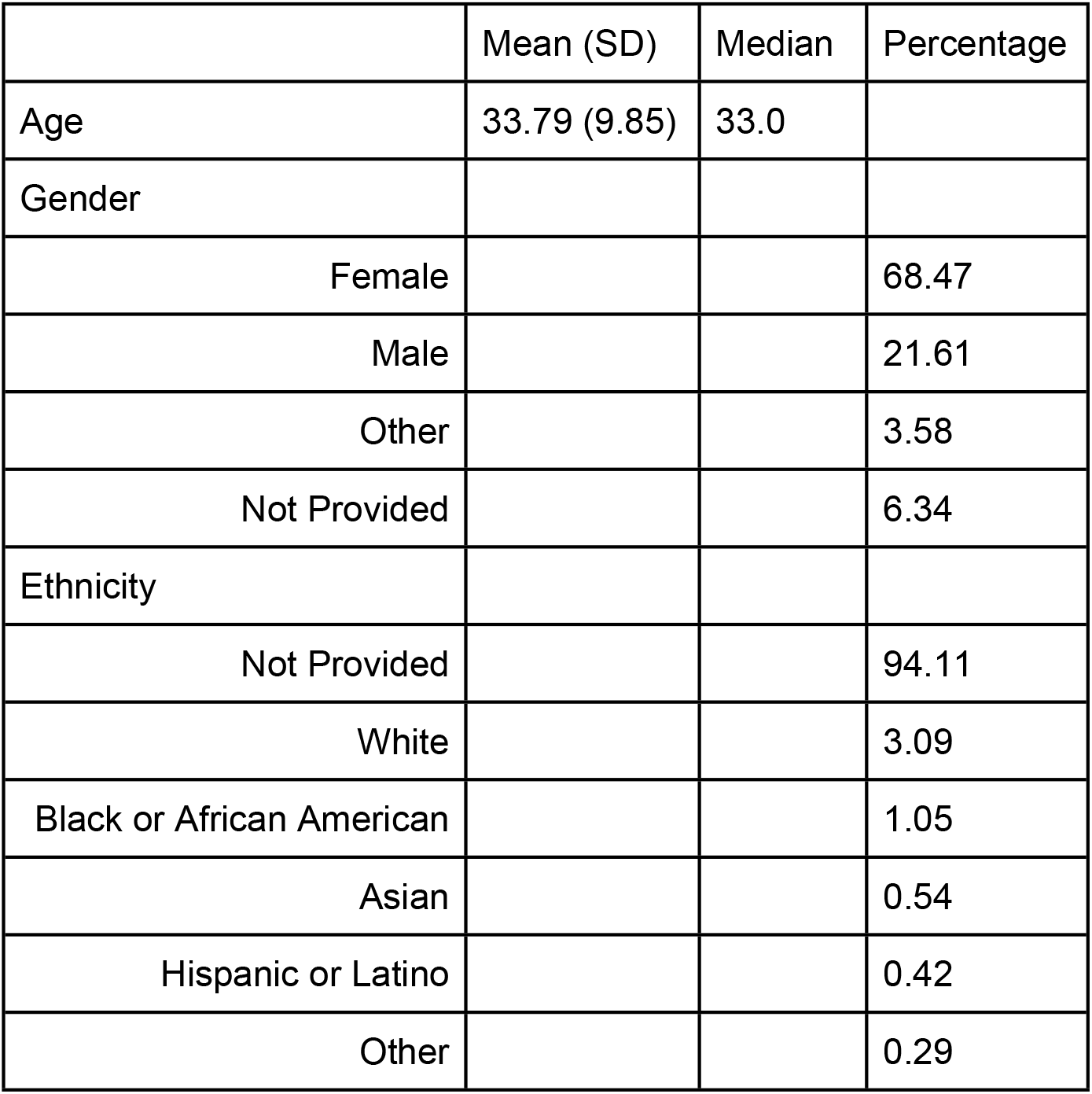
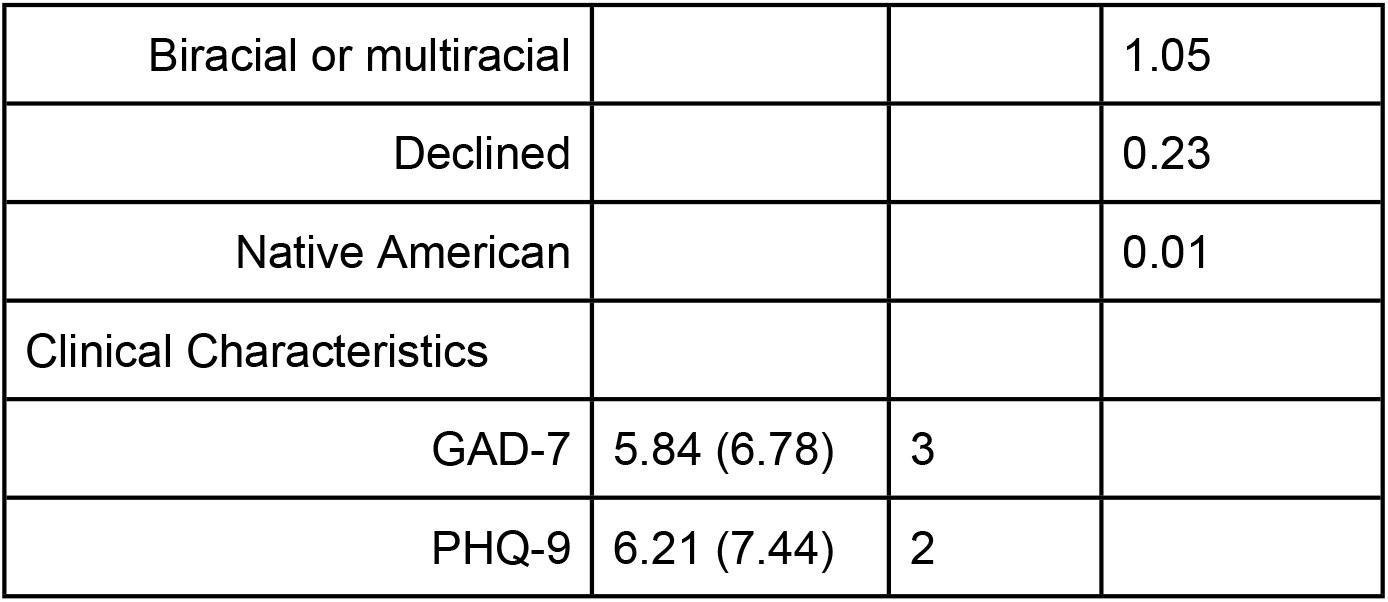

|  | Mean (SD) | Median | Percentage |
| --- | --- | --- | --- |
| Age | 33.79 (9.85) | 33.0 |  |
| Gender |  |  |  |
| Female |  |  | 68.47 |
| Male |  |  | 21.61 |
| Other |  |  | 3.58 |
| Not Provided |  |  | 6.34 |
| Ethnicity |  |  |  |
| Not Provided |  |  | 94.11 |
| White |  |  | 3.09 |
| Black or African American |  |  | 1.05 |
| Asian |  |  | 0.54 |
| Hispanic or Latino |  |  | 0.42 |
| Other |  |  | 0.29 |
| Biracial or multiracial |  |  | 1.05 |
| Declined |  |  | 0.23 |
| Native American |  |  | 0.01 |
| Clinical Characteristics |  |  |  |
| GAD-7 | 5.84 (6.78) | 3 |  |
| PHQ-9 | 6.21 (7.44) | 2 |  |

### LLM as Labeler

The majority of labelled messages were categorized as moderate risk (SI-3: Suicidal Ideation without plan or means; Figure 1). Agreement between the LLM and human consensus achieved a Precision of 0.69, Recall of 0.51, F1 of 0.58 and a weighted F1 score of 0.88.

**Figure 1.**
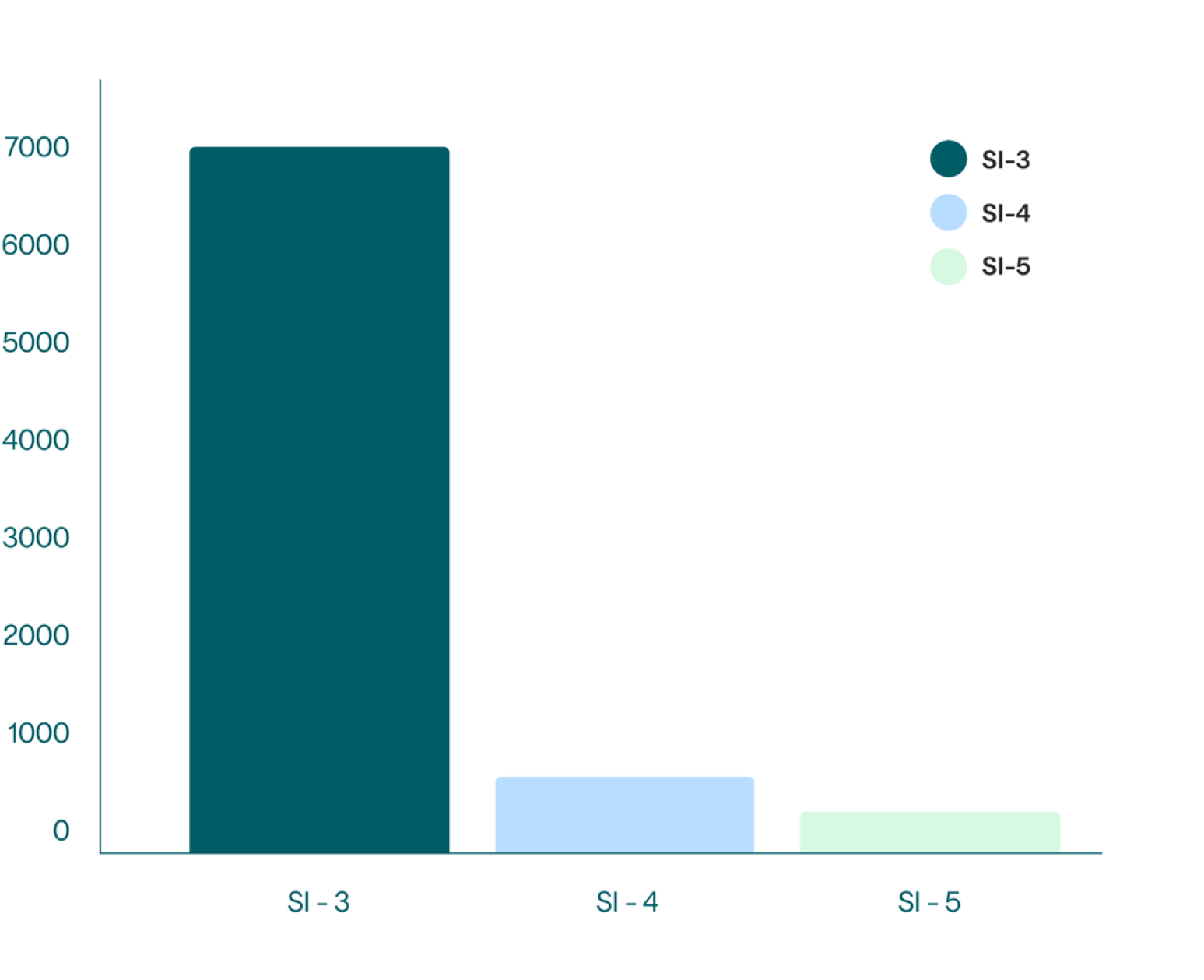
Number of labeled messages in each category.

### Model v2.0

Several modeling approaches were evaluated, using both embeddings from transformer models including RoBERTa^12^ and ELECTRA,^13^ and extracted features from the text corpus (Table 4). The best performing model was a fine-tuned RoBERTa foundation model (Figure 2).

**Table 4.** Evaluation metrics for SI model v2.0.

| Table 4. Evaluation metrics for SI model v2.0 |  |  |  |  |  |  |
| --- | --- | --- | --- | --- | --- | --- |
| Model | F1 | F2 | Precision | Recall | AUC | Accuracy |
| LR | 0.72 | 0.65 | 0.50 | 0.71 | 0.72 | 0.71 |
| NB | 0.71 | 0.62 | 0.51 | 0.66 | 0.77 | 0.72 |
| KNN | 0.73 | 0.64 | 0.51 | 0.69 | 0.77 | 0.72 |
| RF | 0.77 | 0.67 | 0.57 | 0.70 | 0.81 | 0.76 |
| XGB | 0.80 | 0.66 | 0.65 | 0.66 | 0.83 | 0.80 |
| 3 layer NN | 0.71 | 0.58 | 0.49 | 0.64 | 0.74 | 0.71 |
| **RoBERTa** | **0.90** | **0.81** | **0.83** | **0.81** | **0.94** | **0.93** |
| ELECTRA | 0.89 | 0.79 | 0.81 | 0.79 | 0.93 | 0.89 |

**Figure 2.**
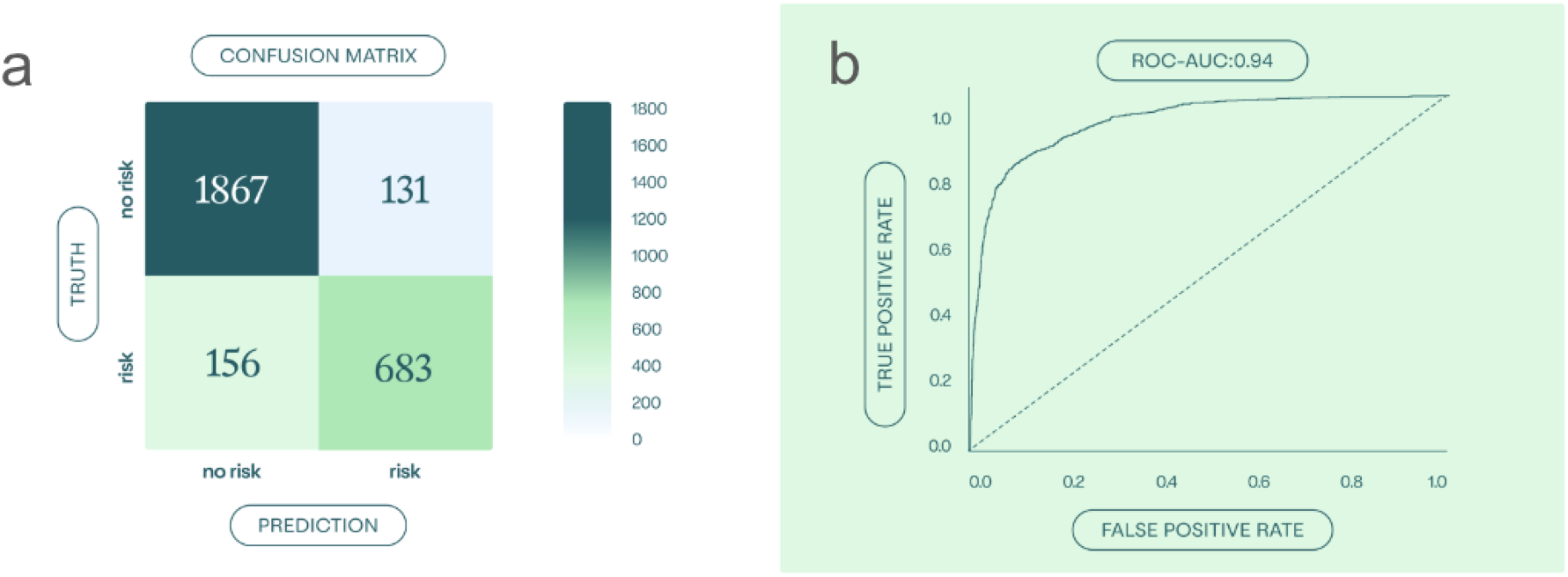
a. Confusion matrix for model v2.0 for the best performing model, a fine-tuned RoBERTa model trained to classify messages as “risky” or “not risky” b. ROC-AUC curve for the RoBERTa binary classification model, AUC = 0.94.

**Figure 3.**
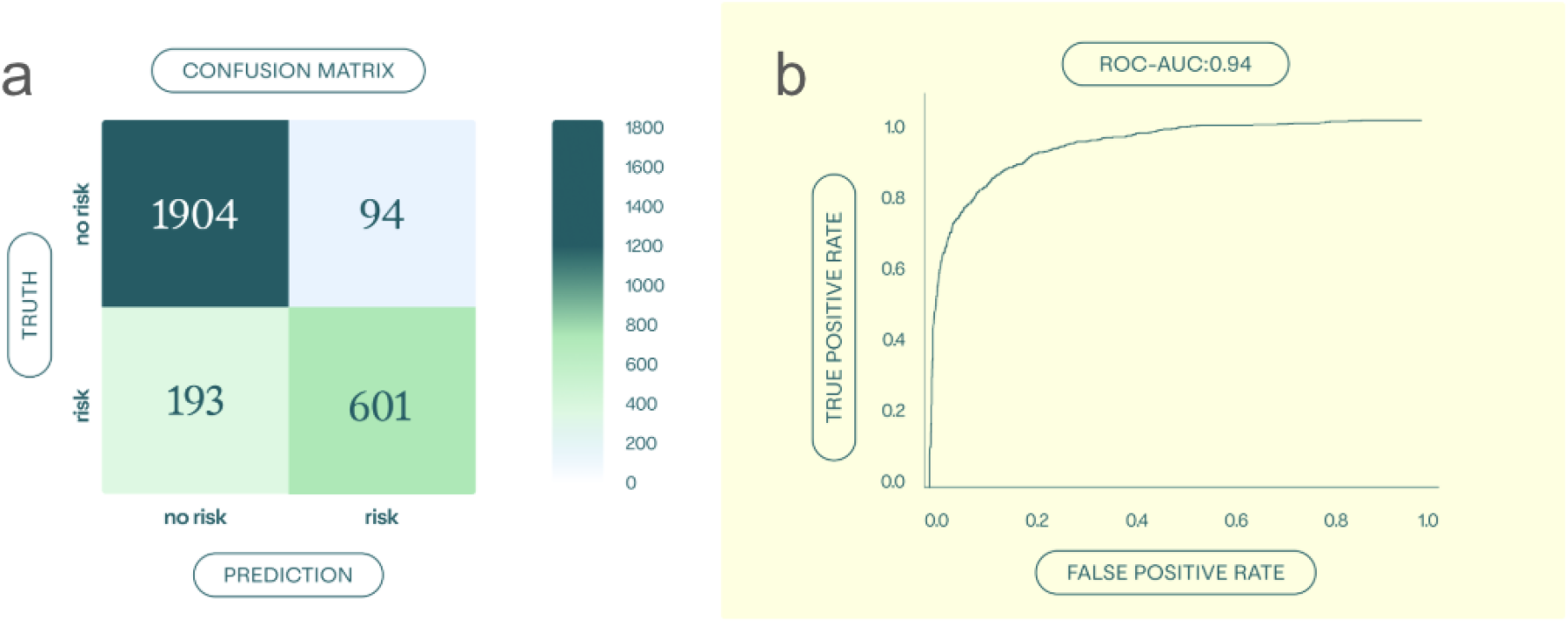
a. Confusion matrix for model v2.1 for the best performing model, and Autogluon model trained to classify messages “risky” or “not risky” b. ROC-AUC curve for the RoBERTa binary classification model, AUC = 0.94.

### Model v2.1

The addition of demographic and census-derived SDOH metrics did increase some of the accuracy metrics for the model, namely increased precision, but also increased the number of risky messages the model missed (false negatives). This model was trained using Autogluon multimodal^25^ which attempts a number of encoding and modeling architectures, and determines the best fitting model (Table 5).

**Table 5.**
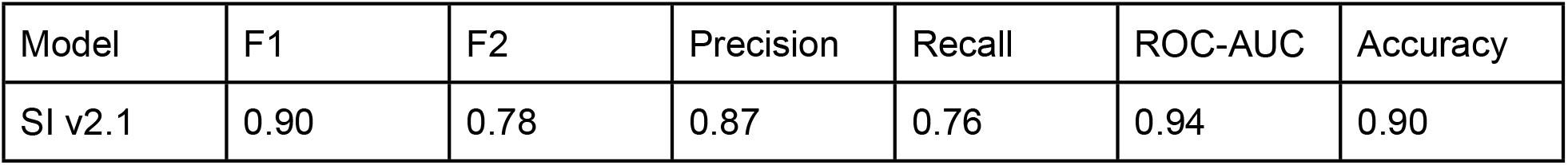
Evaluation metrics for SI model v2.1.

To assess whether any of these additional groups of features has an outsized influence on the performance of the model, we also trained three additional models where we withheld either census derived SDOH data, demographics, or survey information. Removing each group led to very slight decreases across the different accuracy metrics, but no substantial changes were seen in any specific group (Table 6; Figure 4).

**Table 6.** Evaluation metrics for SI model v2.1 when each variable was left out.

| Left Out | F1 | F2 | Precision | Recall | AUC | Accuracy |
| --- | --- | --- | --- | --- | --- | --- |
| Census Data | .89 | .78 | .85 | .76 | .94 | .89 |
| Demographics | .88 | .77 | .81 | .76 | .93 | .88 |
| Surveys | .89 | .81 | .79 | .82 | .94 | .89 |

**Figure 4.**
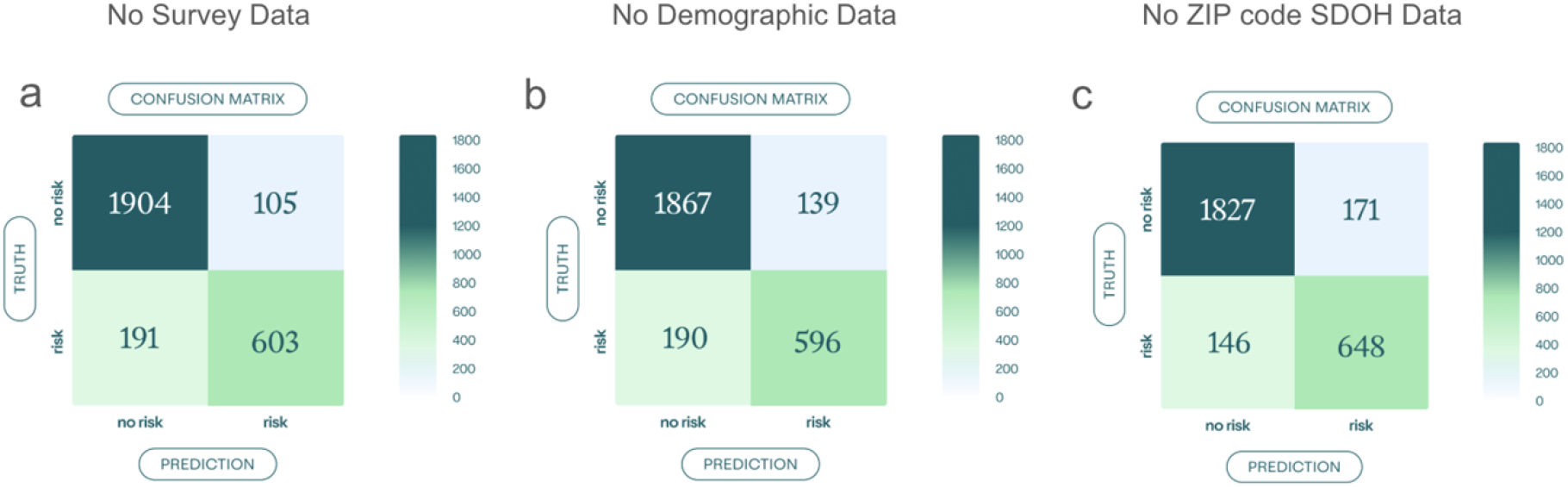
Confusion matrices for model v2.1 left out models a) no survey data, b) no demographic data, c) no ZIP code level SDOH data.

Because the addition of SDOH data, demographics, and survey information did not have a major impact on F1 or F2 scores, and led to decreased recall, they were not included in subsequent models.

### Model v3.0

The final iteration implemented two multiclass models using embeddings from RoBERTa^12^ and ELECTRA.^13^ While model v3.0’s F1 and F2 scores declined slightly relative to its binary predecessor (v2.1), it was still effective in critical scenarios (Table 7 and Figure 5). In this case, only the RoBERTa fine-tuned sequence classification algorithms could discriminate between all three groups. When the RoBERTa model misclassified severe risk, these misclassifications were typically labeled as “moderate,” and rarely (n = 7/76) labeled “no risk.” The fine-tuned ELECTRA sequence classification models could not discriminate between moderate and severe risk.

**Table 7.** Evaluation metrics for SI model v3.0.

| *RoBERTa* |  | F1 | F2 | precision | recall |
| --- | --- | --- | --- | --- | --- |
|  | no risk | 0.89 | 0.89 | 0.90 | 0.89 |
|  | moderate | 0.82 | 0.82 | 0.83 | 0.82 |
|  | severe | 0.49 | 0.52 | 0.45 | 0.54 |
|  | macro avg | 0.73 | 0.75 | 0.72 | 0.75 |
|  | weighted avg | 0.85 |  | 0.85 | 0.85 |
|  | accuracy | 0.85 |  |  |  |
| *ELECTRA* | no risk | 0.92 | 0.92 | 0.93 | 0.90 |
|  | moderate | 0.75 | 0.79 | 0.70 | 0.82 |
|  | severe | 0.00 | 0 | 0.00 | 0.00 |
|  | macro avg | 0.55 | 0.57 | 0.54 | 0.57 |
|  | weighted avg | 0.85 |  | 0.84 | 0.86 |
|  | accuracy | 0.86 |  |  |  |

**Figure 5.**
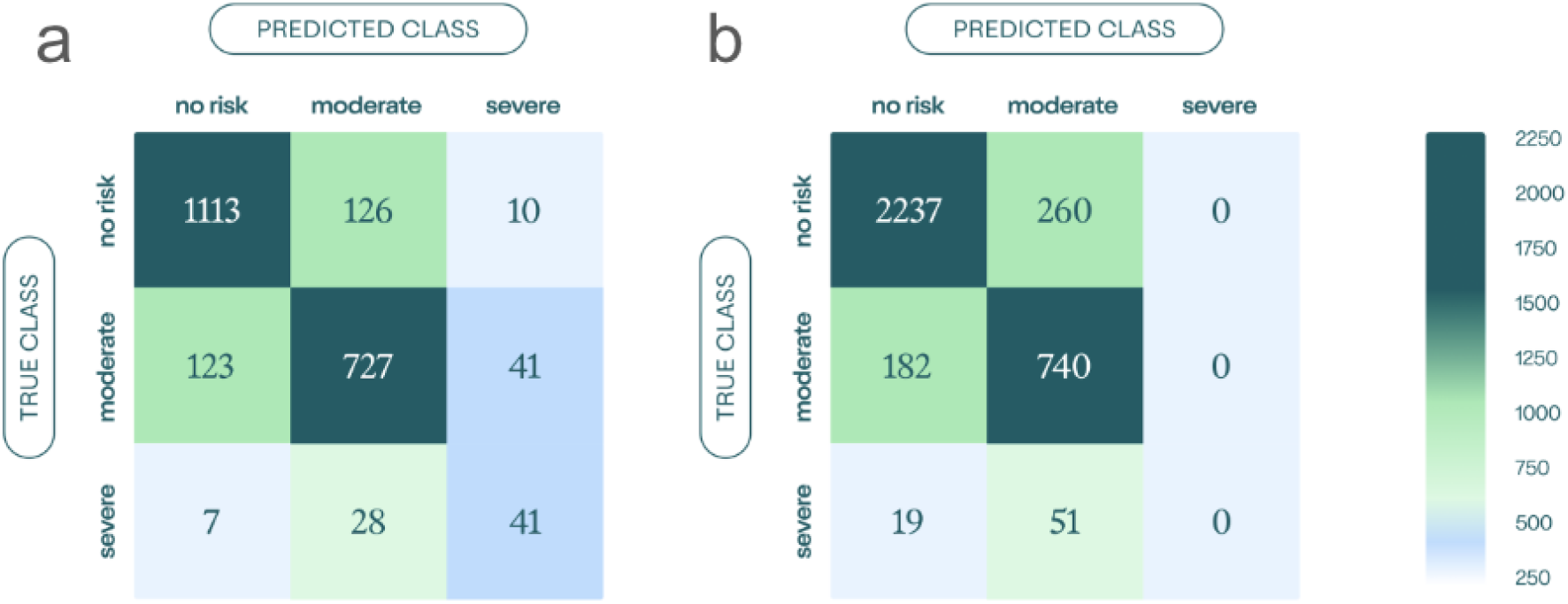
Confusion matrix and evaluation metrics for SI model v3.0 a) Multiclass RoBERTa fine-tuned model b) Multiclass ELECTRA fine-tuned model

## Discussion

### Key Results

This study demonstrates that modern machine learning techniques can measurably enhance the detection of suicidal ideation (SI) risk within real-world clinical transcripts. All three models developed in this study outperform the previous SI model across all accuracy metrics, particularly in terms of precision. The current models have a weighted F1-score of > 0.84 compared to an F1 of 0.18 for the original production model.^5^

The most clinically significant finding is the improvement in precision, which rose from 0.11 in earlier iterations to over 0.75 in the current study. This reduction in “false alarms” ensures that clinicians can focus their resources on genuine risk without the burden of excessive, non-actionable alerts. Simultaneously, these models maintain high sensitivity, capturing a greater proportion of actual risk articulated in asynchronous therapeutic conversations.

The integration of demographic and social determinants of health (SDOH) in version 2.1 was theoretically motivated by the fact that that suicide risk and the linguistic expression of distress (such as communication styles and disclosure patters) are shaped by factors such as age gender, and socioeconomic environment.^26,27^ Cultural norms significantly influence whether a client conveys suicidal ideation (SI) directly or through filtered, indirect language.^27^ However, the finding that these variables provided only marginal utility suggests that these factors do not significantly add to the models’ ability to predict risk, or that our data is not granular enough in these features, which is likely the case, given that many factors are derived at the ZIP code level and not the personal level, and that many of our participants choose not to provide demographic data where it is optional. Thus, we decided to exclude SDOH and survey data from the multiclass model (v3.0).

The transition to the multiclass model (v3.0) was driven by provider requests for more granular risk information to allow for more effective triage. Although the F1 and F2 scores decreased slightly when moving from binary (v2.0, 2.1) to multiclass models, likely due to the added complexity of differentiating between more classes and a lower probability for any single correct guess, the clinical utility of the system was enhanced. The tiered system classifies risk into moderate and severe categories, allowing for an efficient triage process. In clinical practice, this allows providers to prioritize high-acuity cases (those involving methods or plans) for immediate escalation, while managing moderate ideation within the standard therapeutic flow. This stratification can help reduce the phenomenon of alert fatigue, where clinicians may become desensitized to automated warnings, high volumes of alerts, frequent false positives, and a lack of clear prioritization, which can lead to frustration, cognitive overload, and general desensitization over time.^4, 28-29^ By providing severity levels, the system balances patient safety with the cognitive load and usability requirements of the treating provider.^29^

### Limitations

Despite the significant advancements demonstrated by these models, a few limitations are worth noting. First, the models are trained on data from a single digital mental health platform, which may limit generalizability to other clinical settings or communication modalities outside of asynchronous text-based therapy. Future work should expand upon these aims to predict risk from settings outside of therapeutic settings (e.g. everyday messages shared with peers or online). Second, the demographic data was subject to missing values. Had there been more available and nuanced data, we may have observed differences in the model evaluation metrics associated with incorporating that data in v2.1. Finally, the reliance on an LLM-based labeling system, even with human consensus review, may introduce subtle biases or inaccuracies in the ground truth, particularly for the most nuanced expressions of severe risk.

### Conclusions

In conclusion, this work provides a robust, clinically actionable tool that strengthens the digital safety net for clients experiencing suicidal distress while enhancing the efficiency of clinical safety workflows. This new risk-detection system, specifically model v3.0, achieves this by successfully developing and validating a tiered, high-precision machine learning approach for real-time suicide risk classification in digital psychotherapy transcripts, which represents a significant advance over previously deployed production models. Critically, the multiclass nature of the system effectively distinguishes between “no risk,” “moderate ideation,” and “severe risk,” thereby transforming clinical utility by enabling prioritized triage and the optimization of intervention resources to deliver superior care to those in crisis.

## Data Availability

Data cannot be shared publicly because they were collected as part of standard business operations. Data are available from Talkspace for researchers who meet the criteria for access to confidential data.

## Author Contributions

**Macayla L. Donegan:** Conceptualization, Formal analysis, Investigation, Methodology, Software, Validation, Visualization, Writing – original draft, Writing – review & editing, **Agrima Srivastava:** Conceptualization, Formal analysis, Methodology, Software, Validation, Writing –original draft, Writing – review & editing, **Emily Peake:** Project administration, Resources, Writing – original draft, Writing – review & editing, **Mackenzie Swirbul:** Project administration, Resources, Writing – original draft, Writing – review & editing, **Alby Ungashe:** Data curation, Writing – original draft, **Michael J. Rodio:** Conceptualization, Data curation, Project administration, Resources, Writing – review & editing **Nir Tal**: Supervision, **Gil Margolin**: Supervision, **Nikole Benders-Hadi**: Supervision, **Aarthi Padmanabhan:** Conceptualization, Data curation, Project administration, Resources, Writing – original draft, Writing – review & editing

## Acknowledgements

Thank you to Alivya Barry, Kirstin Battles, Devin Nelson, Sheryl Nelson, Mica Poerio, Annika Toivonen, LaShae Williams for their help labeling data. Thank you to Mary Potter (data security and HIPAA compliance), Jack Miller and Brett Jannusch (development and maintenance of the data labeling system) and Jessica Pegram (accounting and financial management).

## References

1. Centers for Disease Control and Prevention. Suicide data and statistics [Internet]. Atlanta (GA): CDC; 2025 [cited 2026 Mar 26]. Available from: https://www.cdc.gov/suicide/facts/data.html

2. Smith M. Suicide risk assessments: a scientific and ethical critique. Bioethical Inq. 2022;19(3):481–93. doi: 10.1007/s11673-022-10189-5.

3. Linthicum KP, Schafer KM, Ribeiro JD. Machine learning in suicide science: applications and ethics. Behav Sci Law. 2019;37(3):214–22. doi: 10.1002/bsl.2392.

4. Agency for Healthcare Research and Quality (AHRQ). Alert fatigue [Internet]. Rockville (MD): Patient Safety Network; 2024 [cited 2026 Mar 26]. Available from: https://psnet.ahrq.gov/primer/alert-fatigue

5. Bantilan N, Malgaroli M, Ray B, Hull TD. Just in time crisis response: suicide alert system for telemedicine psychotherapy settings. Psychother Res. 2021;31(3):289–99. doi: 10.1080/10503307.2020.1842311.

6. Gaur V, Maggu G, Bairwa K, Chaudhury S, Dhamija S, Ali T. Artificial intelligence in suicide prevention: utilizing deep learning approach for early detection. Ind Psychiatry J. 2024;33(2):226–33. doi: 10.4103/ipj.ipj_20_24.

7. Lazaridou A, Kuncoro A, Gribovskaya E, Agrawal D, Liska A, Terzi T, et al. Mind the gap: assessing temporal generalization in neural language models. Adv Neural Inf Process Syst. 2021;34:29348–63.

8. Guo LL, Pfohl SR, Fries J, Posada J, Fleming SL, Aftandilian C, et al. Systematic review of approaches to preserve machine learning performance in the presence of temporal dataset shift in clinical medicine. Appl Clin Inform. 2021;12(4):808–15. doi: 10.1055/s-0041-1735165.

9. Kumar V, Sznajder KK, Kumara S. Machine learning based suicide prediction and development of suicide vulnerability index for U.S. counties. npj Ment Health Res. 2022;1(1):3. doi: 10.1038/s44184-022-00002-x.

10. Xiao Y, Meng Y, Brown TT, Tsai AC, Snowden LR, Chow JC, et al. Machine learning to investigate policy-relevant social determinants of health and suicide rates in the United States. Nat Ment Health. 2025;3:675–84. doi: 10.1038/s44220-025-00424-4.

11. Lopes IM, Seabra D, Santos G, Ramalho N, Coelho Rocha T, Alves Leal J, et al. Beyond self-report: addressing suicide risk in personal narrative crisis. Eur Psychiatry. 2025;68(S1):S1166–7. doi: 10.1192/j.eurpsy.2025.2361.

12. Liu Y, Ott M, Goyal N, Du J, Joshi M, Chen D, et al. RoBERTa: a robustly optimized BERT pretraining approach. arXiv [Preprint]. 2019. 1907.11692.

13. Clark K, Luong MT, L. QV, Manning CD. ELECTRA: Pre-training text encoders as discriminators rather than generators. arXiv [Preprint]. 2020. 2003.10555.

14. Pedregosa F, Varoquaux G, Gramfort A, Michel V, Thirion B, Grisel O, et al. Scikit-learn: machine learning in Python. J Mach Learn Res. 2011;12:2825–30.

15. Akiba T, Sano S, Yanase T, Ohta T, Koyama M. Optuna: a next-generation hyperparameter optimization framework. In: Proceedings of the 25th ACM SIGKDD International Conference on Knowledge Discovery & Data Mining; 2019 Jul 25; Anchorage, AK. New York: ACM; 2019. p. p2623-31. doi: 10.1145/3292500.3330701.

16. Sacco SJ, Chen K, Wang F, Aseltine R. Target-based fusion using social determinants of health to enhance suicide prediction with electronic health records. PLoS One. 2023;18(4):e0283595. doi: 10.1371/journal.pone.0283595.

17. Perry SW, Rainey JC, Allison S, Bastiampillai T, Wong ML, Licinio J, et al. Achieving health equity in US suicides: a narrative review and commentary. BMC Public Health. 2022;22(1):1360. doi: 10.1186/s12889-022-13596-w.

18. Carmichael AE, Lennon NH, Qualters JR. Analysis of social determinants of health and individual factors found in health equity frameworks: applications to injury research. J Safety Res. 2023;87:508–18. doi: 10.1016/j.jsr.2023.00.001.

19. Alemi F, Avramovic S, Renshaw KD, Kanchi R, Schwartz M. Relative accuracy of social and medical determinants of suicide in electronic health records. Health Serv Res. 2020;55 Suppl 2:833–40. doi: 10.1111/1475-6773.03540.

20. Kirkbride JB, Anglin DM, Colman I, Dykxhoorn J, Jones PB, Patalay P, et al. The social determinants of mental health and disorder: evidence, prevention and recommendations. World Psychiatry. 2024;23(1):58–90. doi: 10.1002/wps.21160.

21. Monnat SM. Trends in U.S. working-age non-Hispanic white mortality: rural-urban and within-rural differences. Popul Res Policy Rev. 2020;39(5):805–34. doi: 10.1007/s11113-020-09607-6.

22. Stein EM, Gennuso KP, Ugboaja DC, Remington PL. The epidemic of despair among white Americans: trends in the leading causes of premature death, 1999-2015. Am J Public Health. 2017;107(10):1541–7. doi: 10.2105/AJPH.2017.303941.

23. Case A, Deaton A. Rising morbidity and mortality in midlife among white non-Hispanic Americans in the 21st century. Proc Natl Acad Sci U S A. 2015;112(49):15078–83. doi: 10.1073/pnas.1518393112.

24. Kind AJH, Buckingham WR. Making neighborhood-disadvantage metrics accessible -the Neighborhood Atlas. N Engl J Med. 2018;378(26):2456–8. doi: 10.1056/NEJMp1802313.

25. Tang Z, Shao B, Chou JC, Mueller J, Smola A, Erickson N. AutoGluon-multimodal (AutoMM): supercharging multimodal AutoML with foundation models. arXiv [Preprint]. 2024. 2404.16233.

26. Feldhege J, Wolf M, Moessner M, Bauer S. Psycholinguistic changes in the communication of adolescent users in a suicidal ideation online community during the COVID-19 pandemic. Eur Child Adolesc Psychiatry. 2023;32:975–85. doi: 10.1007/s00787-022-02067-7.

27. Rodriguez TR, Bandel SL, Daruwala SE, Anestis MD, Anestis JC. Predictors and patterns of suicidal ideation disclosures among American adults. Suicide Life Threat Behav. 2025;55:e13126. doi: 10.1111/sltb.13126.

28. Khairat S, Burke G, Archambault H, Schwartz T, Larson J, Ratwani R. Perceived burden of EHR-based alerts and warnings by clinicians: a systematic review. Health Informatics J. 2021;27(3):1–12. doi: 10.1177/1460458221100753.

29. Gordon WJ, Khurshid A, Landman AB. Reducing alert fatigue in clinical decision support systems: a review of best practices and implementation strategies. Digit Health. 2023;9:1–10. doi: 10.1016/j.dhjo.2023.001159.

